# Temporal increase in D614G mutation of SARS-CoV-2 in the Middle East and North Africa: Phylogenetic and mutation analysis study

**DOI:** 10.1101/2020.08.24.20176792

**Authors:** Malik Sallam, Nidaa A. Ababneh, Deema Dababseh, Faris G. Bakri, Azmi Mahafzah

## Abstract

Phylogeny construction can help to reveal evolutionary relatedness among molecular sequences. The spike (*S*) gene of SARS-CoV-2 is the subject of an immune selective pressure which increases the variability in such region. This study aimed to identify mutations in the *S* gene among SARS-CoV-2 sequences collected in the Middle East and North Africa (MENA), focusing on the D614G mutation, that has a presumed fitness advantage. Another aim was to analyze the *S* gene sequences phylogenetically. The SARS-CoV-2 *S* gene sequences collected in the MENA were retrieved from the GISAID public database, together with its metadata. Mutation analysis was conducted in Molecular Evolutionary Genetics Analysis software. Phylogenetic analysis was done using maximum likelihood (ML) and Bayesian methods. A total of 553 MENA sequences were analyzed and the most frequent *S* gene mutations included: D614G = 435, Q677H = 8, and V6F = 5. A significant increase in the proportion of D614G was noticed from (63.0%) in February 2020, to (98.5%) in June 2020 (p< 0.001). Two large phylogenetic clusters were identified via ML analysis, which showed an evidence of inter-country mixing of sequences, which dated back to February 8, 2020 and March 15, 2020 (median estimates). The mean evolutionary rate for SARS-CoV-2 was about 6.5 × 10^−3^ substitutions/site/year based on large clusters’ Bayesian analyses. The D614G mutation appeared to be taking over the COVID-19 infections in the MENA. Bayesian analysis suggested that SARS-CoV-2 might have been circulating in MENA earlier than previously reported.

## Introduction

Members of *Coronaviridae* family of viruses have started to gain a substantial interest due to their potential role as causative agents of emerging infections in humans (Fehr and Perlman, 2015). This was manifested by the 2002–2003 SARS outbreak, 2012 MERS outbreak, and the current coronavirus disease 2019 (COVID-19) pandemic, the first documented coronavirus pandemic, which can be viewed as the full-blown consequence of coronavirus threat (Cherry and Krogstad, 2004; Liu *et al*., 2020; Lu and Liu, 2012; Peiris *et al*., 2003).

The causative agent of this unprecedented pandemic is an enveloped RNA virus of the subfamily *Betacoronavirinae* (Liu *et al*., 2020). Similar to other RNA viruses, severe acute respiratory syndrome coronavirus 2 (SARS-CoV-2), is presumed to have a relatively high mutation rate that is mostly related to its RNA-dependent RNA polymerase, with minimal proofreading activity (Duffy *et al*., 2008; Sevajol *et al*., 2014). In addition, the high frequency of recombination in coronaviruses augments its genetic diversity and its ability of cross-species transmission (Su *et al*., 2016; Woo *et al*., 2009).

The aforementioned features are accompanied by ubiquitous presence of coronaviruses in various animal reservoirs (Guan *et al*., 2003). Thus, cross-species transmission, including spread to humans seems an inevitable outcome (Graham and Baric, 2010; Woo *et al*., 2009). This is mainly related to human, ecologic and economic factors, which explain the increased frequency of zoonosis (Delabouglise *et al*., 2017; Karesh *et al*., 2012; Morse *et al*., 2012).

The transcripts of SARS-CoV-2 include nine sub-genomic RNAs, of which one structural protein, spike glycoprotein, being responsible for attachment of the virus to its cellular receptor (angiotensin-converting enzyme 2 [ACE2]) (Fehr and Perlman, 2015). Host proteases’ cleavage of the spike glycoprotein is essential for virion entry into the target cells (Ou *et al*., 2020). The receptor-binding domain (RBD) in the S1 subunit binds ACE2 and facilitates fusion with host cell membrane (Tai *et al*., 2020). For the S2 domain of the spike glycoprotein, its function facilitates fusion of the viral and host cell membranes (Xia *et al*., 2020).

Studying the SARS-CoV-2 *S* gene attracts a special attention, particularly from an immunologic and evolutionary points of view (Chen *et al*., 2020; Korber *et al*., 2020; Robson, 2020). The spike gene is the subject of an immune selective pressure, and antibodies against its protein product can inhibit the viral entry into the target cells (Korber *et al*., 2020; Walls *et al*., 2020). The selective forces directed against the *S* gene can increase genetic variability in the region, which can be used to infer the evolutionary relationships between viral sequences in a shorter time, compared to use of less variable regions (e.g. RNA-dependent RNA polymerase gene (*RdRp*)), where mutations occur, but appear to be more costly (Duffy *et al*., 2008; Moya *et al*., 2004; Pachetti *et al*., 2020; Robson, 2020).

Genetic variability in the *S* gene can be demonstrated by continuous emergence of mutations, that were reported at a global level (Korber *et al*., 2020). Some of these mutations appeared to have a significant epidemiologic value, with the replacement of aspartic acid by glycine at position 614 of the spike glycoprotein (D614G), which is associated with a higher viral shedding and increased infectivity (Korber *et al*., 2020; Maitra *et al*., 2020; Zhang *et al*., 2020). This mutation currently appears to be dominating the pandemic (Grubaugh *et al*., 2020). However, the clinical effect of such mutation is yet to be fully determined (Eaaswarkhanth *et al*., 2020; Kim *et al*., 2020b; Korber *et al*., 2020). Other mutations in the *S* gene have also been reported, with the most frequent including: D936Y/H, P1263L, and L5F (Korber *et al*., 2020; Lokman *et al*., 2020).

Similar to other RNA viruses, SARS-CoV-2 can be the subject of phylogenetic analysis due to its high evolutionary rate, and the application of molecular clock analysis might be of value to determine the timing of introductions of large clusters that imply networks of transmission (Duffy *et al*., 2008; Forster *et al*., 2020; Pybus and Rambaut, 2009). State-of-the-art methods for phylogeny construction include maximum likelihood and Bayesian tools (Anisimova *et al*., 2013).

The Middle East and North Africa (MENA) region include the following 19 countries: Algeria, Bahrain, Egypt, Iran, Iraq, Jordan, Kuwait, Lebanon, Libya, Morocco, Oman, Palestine, Qatar, Kingdom of Saudi Arabia (KSA), Sudan, Syria, Tunisia, United Arab Emirates (UAE), and Yemen. Countries of the MENA were affected early on during the course of COVID-19 pandemic, with an overwhelming number of cases in some countries (e.g. Iran) (Karamouzian and Madani, 2020; Sawaya *et al*., 2020). The first confirmed cases of COVID-19 in the MENA dated back to February 2020 and were reported in UAE, Iran, and Egypt (Daw *et al*., 2020; Karamouzian and Madani, 2020; Mehtar *et al*., 2020). The total number of diagnosed cases of COVID-19 in the MENA exceeded 1,175,000 with more than 32,000 deaths reported as a result of the disease, as of July 25, 2020 (Worldometer, 2020).

Special attention to COVID-19 infections is needed in the countries of the MENA region, where political and economic factors might lead to devastating effects on the countries affected by the current pandemic (Karamouzian and Madani, 2020; Sawaya *et al*., 2020). Particular attention should be paid to countries like Yemen, Syria and Libya, where the ongoing instabilities can result in underreporting of COVID-19 cases and heavy burden on their health-care systems (Da’ar *et al*., 2020; Daw, 2020; Karamouzian and Madani, 2020; Sawaya *et al*., 2020).

The aims of this study included an attempt to phylogenetically analyze *S* gene sequences and to analyze the spike gene mutation patterns in the MENA region. In addition, we aimed to characterize the temporal changes of D614G mutation spread in the region.

## Materials and Methods

### Compilation of the MENA SARS-CoV-2 dataset

All SARS-CoV-2 sequences from the MENA countries, were retrieved from the global science initiative and primary source for genomic data of influenza viruses (GISAID) (Elbe and Buckland-Merrett, 2017). We also downloaded the following sequence metadata if available: date of sequence collection, age, gender, city of collection together with country of sequence collection. The sequences were then aligned to the reference SARS-CoV-2 sequence (accession number: NC_045512) and alignment was conducted using multiple alignment program for amino acid or nucleotide sequences (MAFFT v.7) (Rozewicki *et al*., 2019). The MENA sequences that did not contain the complete *S* region were filtered out. In addition, we removed the sequences that contained indels, the nucleotide ambiguity (N); while other ambiguities were retained. The sequences that contained stop codons were removed as well. Each sequence header was also edited to include data in the following order: country of collection, collection date in days starting from January 5, 2020 (the date of reference sequence collection), city, accession number, gender, and age. The final dataset included 553 MENA *S* nucleotide sequences that were collected during January 2020 until June 2020.

### Detection of the S gene mutations

Analysis of the full MENA SARS-CoV-2 *S* gene sequences was conducted in Molecular Evolutionary Genetics Analysis software (MEGA6) (Tamura *et al*., 2013). Visual inspection of the aligned MENA amino acid sequences was done, and mutations were identified based on comparison to the reference SARS-CoV-2 sequence (accession number: NC_045512), which was considered as the wild-type. Amino acids that were translated from codons containing ambiguous bases (e.g. R, Y), were excluded from mutation analysis.

### Maximum likelihood phylogenetic analysis

The whole MENA *S* gene dataset was analyzed phylogenetically using the maximum likelihood (ML) approach in PhyML v3, with selection of the best nucleotide substitution model using Smart Model Selection (SMS), and depending on Akaike Information Criterion (AIK) (Guindon and Gascuel, 2003; Lefort *et al*., 2017). The model which yielded the smallest AIC was the general time-reversible plus invariant sites (GTR + I) nucleotide substitution model with an estimated proportion of invariable sites of 0.625. The estimation of nodal support in the ML tree was based on the approximate Likelihood Ratio Test Shimodaira-Hasegawa like (aLRT-SH) with 0.90 as the statistical significance level (Anisimova *et al*., 2011). The ML analysis was repeated ten times and the ML tree with the highest likelihood was retained for final analysis, and determination of the MENA phylogenetic clusters was done by examining the ML tree from root to tips looking for branches with aLRT-SH ≥ 0.90, with large clusters having ≥ 15 sequences.

### Bayesian estimation of time to most recent common ancestors (tMRCAs) of the large MENA phylogenetic clusters

For the large phylogenetic clusters (containing ≥ 15 sequences and identified using ML analysis), tMRCAs were estimated using the Bayesian Markov chain Monte Carlo (MCMC) method implemented in BEAST v1.8.4 (Drummond *et al*., 2012). Bayesian analysis parameters included: Hasegawa–Kishono–Yano (HKY) nucleotide substitution model with discrete gamma-distributed rate heterogeneity, uncorrelated relaxed clock model with a normally-distributed rate prior (initial and mean values of 0.0068, standard deviation = 0.0008), and a Bayesian skyline tree density model (Tang *et al*., 2020). For each large phylogenetic cluster, one run with 200 million chain length was performed. Samples of trees and parameters were collected every 20,000 steps after discarding a burn-in of 20%, and convergence was analyzed in Tracer v1.6.0 (Rambaut *et al*., 2015). The runs were accepted based on effective sample sizes (ESS) of ≥200 and convergence in the trace file. The maximum clade credibility (MCC) trees were assembled using TreeAnnotator in BEAST and were visualized using FigTree (Rambaut, 2012).

### Statistical analysis

Chi-squared test (χ^2^ test) was used to detect differences between the D614 and D614G groups in relation to gender and region (Middle East vs. North Africa). Mann-Whitney *U* test (M-W) was used to assess the difference between the D614 and D614G groups in relation to age. Linear-by-linear test for association (LBL) was used to assess the temporal changes in D614G prevalence. The statistical significance for all aforementioned tests was considered for p< 0.050.

### Sequence accession numbers

A complete list of the MENA SARS-CoV-2 sequence epi accession numbers that were analyzed in this study is provided in (Appendix S1). These sequences are available publicly for registered users of GISAID (Shu and McCauley, 2017).

## Results

### The final MENA SARS-CoV-2 S gene sequence dataset

The total number of MENA SARS-CoV-2 *S* gene sequences that were included in final analysis was 553, distributed as follows: Oman (n = 159), KSA (n = 140), Egypt (n = 95), Morocco (n = 35), Bahrain (n = 34), UAE (n = 32), Jordan (n = 22), Tunisia (n = 8), Kuwait (n = 7), Qatar (n = 7), Lebanon (n = 6), Iran (n = 5), and Algeria (n = 3). The final length of the alignment was 3822 bases. Characteristics of the sequences are highlighted in (Table 1).

**Table 1.**
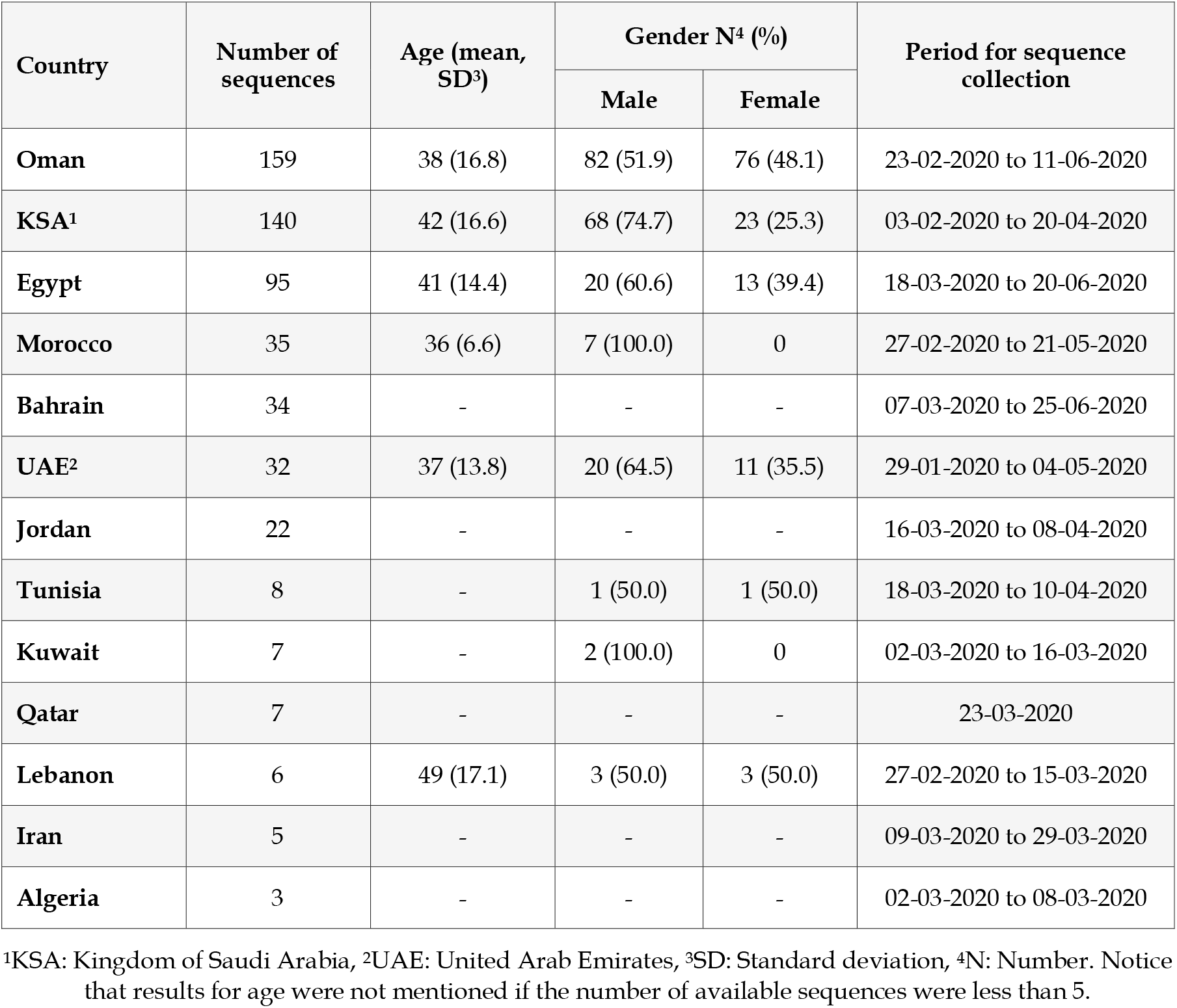
Characteristics of SARS-CoV-2 sequences collected in the Middle East and North Africa and its metadata.

### SARS-CoV-2 *S* gene mutations detected in the MENA

A total 55 unique non-synonymous mutations in the *S* gene were detected as compared to the reference SARS-CoV-2 genome. Eight mutations were identified in spike receptor binding domain (SRD), compared to 21 mutations in S2 glycoprotein domain and 26 in other *S* regions. The most frequent mutation detected in the whole *S* region was D614G (n = 435), followed by Q677H (n = 8), and V6F (n = 5). The majority of mutations were detected sporadically (n = 43, 78.2%, Table 2). The highest number of unique *S* gene mutations (including D614G) was noticed in Oman (n = 16), followed by Egypt (n = 15), Bahrain (n = 9), and KSA (n = 6, Table 2).

**Table 2.**
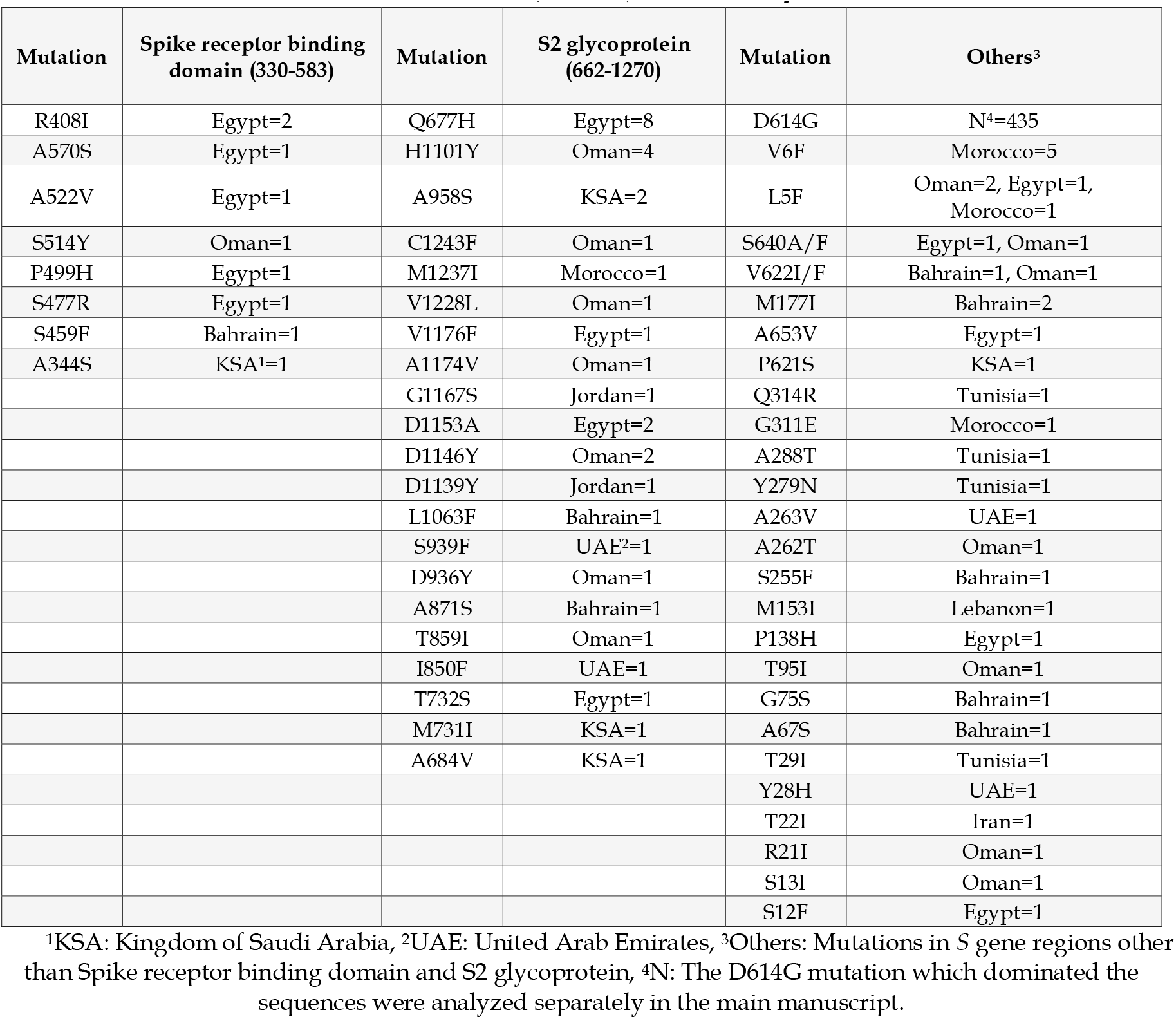
Non-synonymous mutations in the spike (*S*) gene that were detected in the Middle East and North Africa (MENA), stratified by domain.

| Mutation | Spike receptor binding domain (330-583) | Mutation | S2 glycoprotein (662-1270) | Mutation | Others<sup>3</sup> |
| --- | --- | --- | --- | --- | --- |
| R408I | Egypt=2 | Q677H | Egypt=8 | D614G | N<sup>4</sup>=435 |
| A570S | Egypt=1 | H1101Y | Oman=4 | V6F | Morocco=5 |
| A522V | Egypt=1 | A958S | KSA=2 | L5F | Oman=2, Egypt=1, Morocco=1 |
| S514Y | Oman=1 | C1243F | Oman=1 | S640A/F | Egypt=1, Oman=1 |
| P499H | Egypt=1 | M1237I | Morocco=1 | V622I/F | Bahrain=1, Oman=1 |
| S477R | Egypt=1 | V1228L | Oman=1 | M177I | Bahrain=2 |
| S459F | Bahrain=1 | V1176F | Egypt=1 | A653V | Egypt=1 |
| A344S | KSA<sup>1</sup>=1 | A1174V | Oman=1 | P621S | KSA=1 |
|  |  | G1167S | Jordan=1 | Q314R | Tunisia=1 |
|  |  | D1153A | Egypt=2 | G311E | Morocco=1 |
|  |  | D1146Y | Oman=2 | A288T | Tunisia=1 |
|  |  | D1139Y | Jordan=1 | Y279N | Tunisia=1 |
|  |  | L1063F | Bahrain=1 | A263V | UAE=1 |
|  |  | S939F | UAE<sup>2</sup>=1 | A262T | Oman=1 |
|  |  | D936Y | Oman=1 | S255F | Bahrain=1 |
|  |  | A871S | Bahrain=1 | M153I | Lebanon=1 |
|  |  | T859I | Oman=1 | P138H | Egypt=1 |
|  |  | I850F | UAE=1 | T95I | Oman=1 |
|  |  | T732S | Egypt=1 | G75S | Bahrain=1 |
|  |  | M731I | KSA=1 | A67S | Bahrain=1 |
|  |  | A684V | KSA=1 | T29I | Tunisia=1 |
|  |  |  |  | Y28H | UAE=1 |
|  |  |  |  | T22I | Iran=1 |
|  |  |  |  | R21I | Oman=1 |
|  |  |  |  | S13I | Oman=1 |
|  |  |  |  | S12F | Egypt=1 |

<sup>1</sup>KSA: Kingdom of Saudi Arabia, <sup>2</sup>UAE: United Arab Emirates, <sup>3</sup>Others: Mutations in S gene regions other than Spike receptor binding domain and S2 glycoprotein, <sup>4</sup>N: The D614G mutation which dominated the sequences were analyzed separately in the main manuscript.

### Variables associated with a higher prevalence of D614G mutation

Analysis of the two variants of *S* gene (D614 vs. D614G) showed a higher prevalence of D614G in North Africa compared to the Middle East (95.0% vs. 73.7%, p< 0.001; χ^2^ test). In addition, a higher prevalence of D614G variant was noticed in the second half of the study period (April, May and June vs. January, February and March, 90.7% vs. 59.5%, p< 0.001; χ^2^ test). However, no statistical difference was noticed upon comparing the two variants based on age (p = 0.195; M-W), age group (less than 40 years vs. more than or equal to 40 years, p = 0.176; χ^2^ test), or gender (p = 0.644; χ^2^ test). Analysis of the D614G mutant per country showed its presence in all MENA countries included in the study with exception of Iran and Qatar (Figure 1). In addition, no statistical difference was found in analysis per country upon comparing the two variants based on age, age group, or gender.

**Figure 1.**
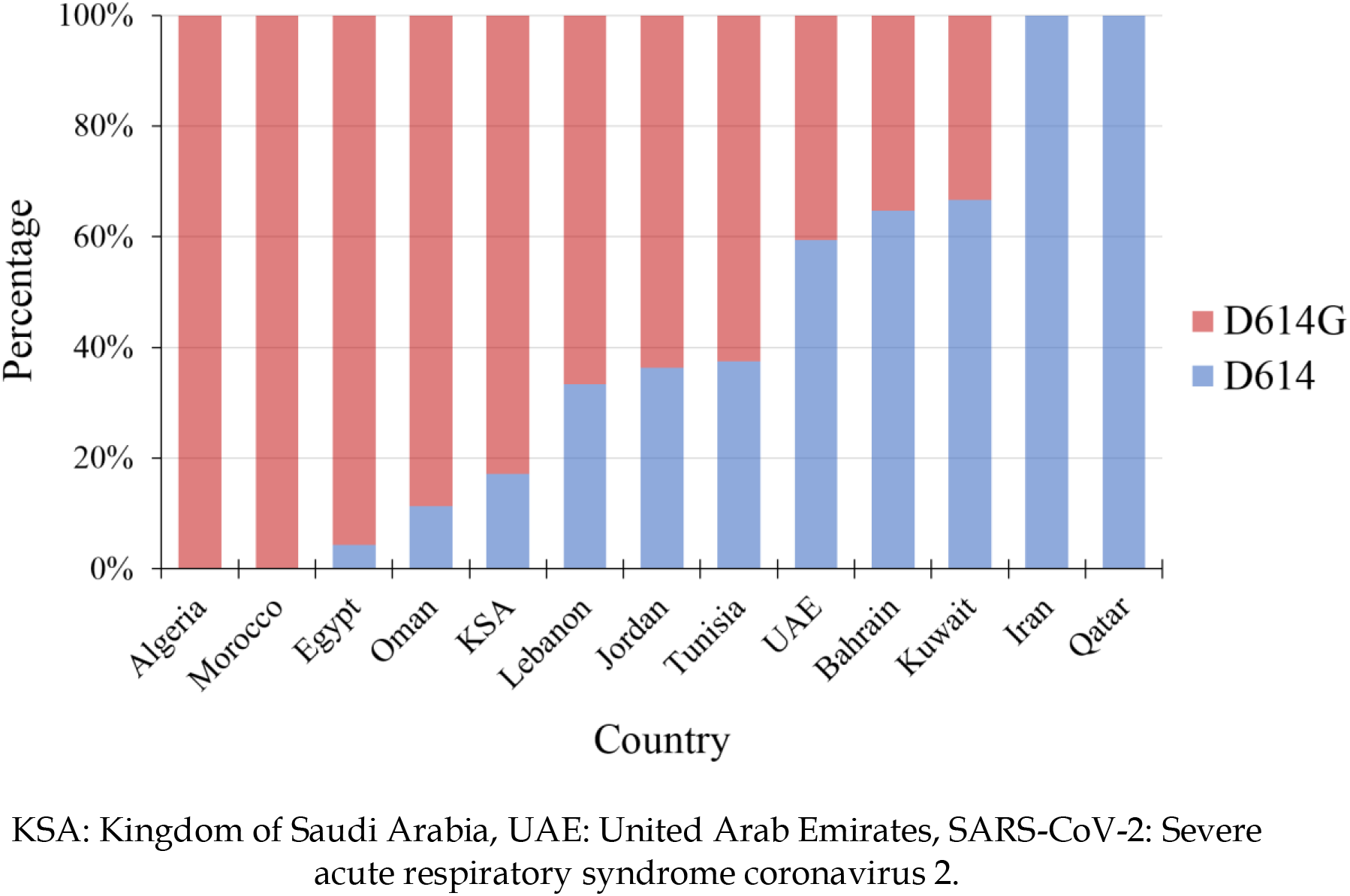
The relative proportions of D614 and D614G mutation in the Middle East and North Africa stratified by countries of SARS-CoV-2 sequence collection.

### Temporal trend of D614G mutant spread in the MENA

Analysis of temporal trend of spread of the D614G mutant of SARS-CoV-2 in the whole MENA region as a single unit revealed an increasing prevalence of D614G from 63.0% in January 2020 to reach 98.5% in June 2020 (p< 0.001; LBL, Figure 2). The same pattern was detected upon comparing the first three months of 2020, compared to April, May and June 2020 (59.5% vs. 90.7%; p< 0.001; χ^2^ test).

**Figure 2.**
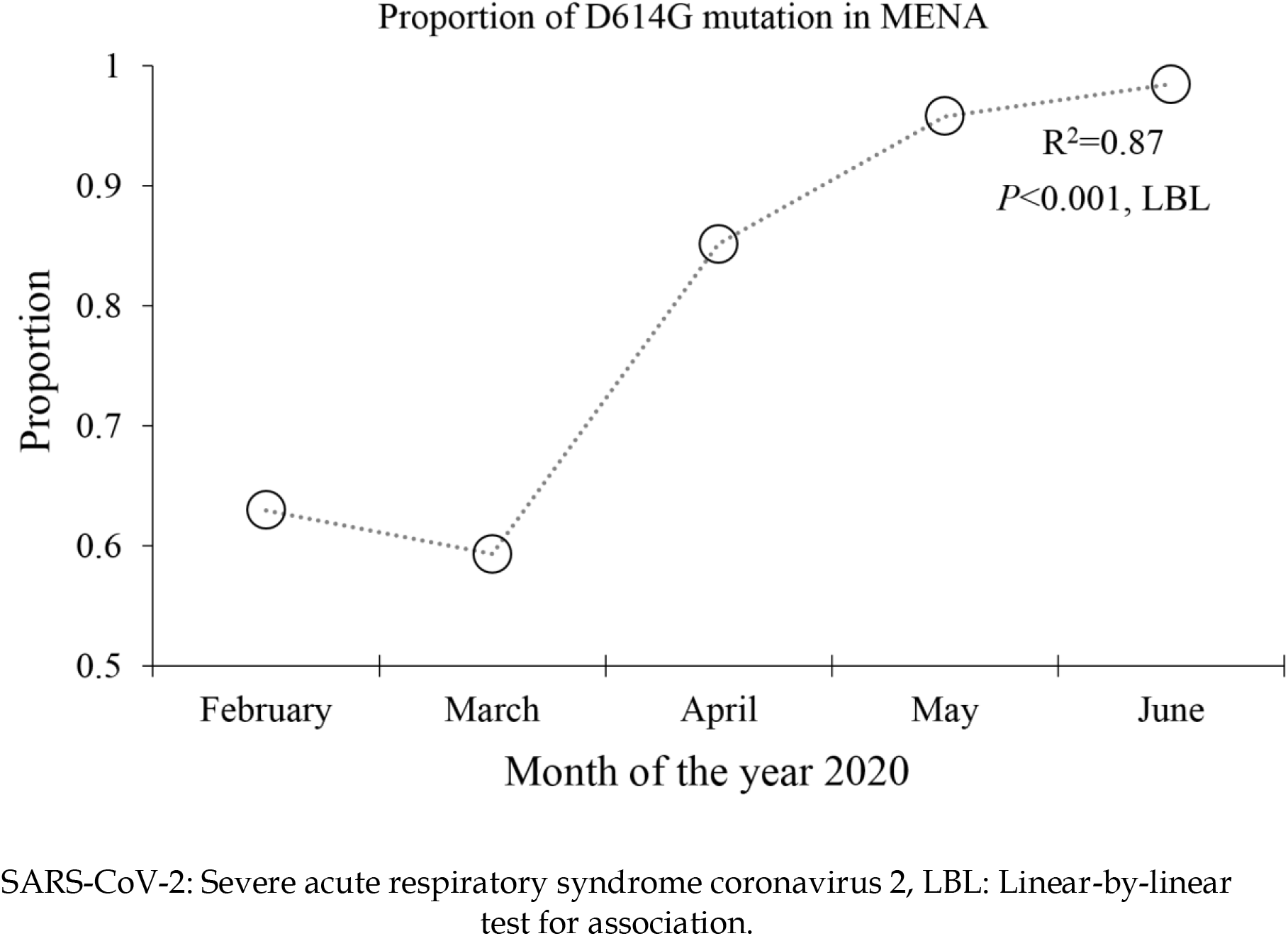
Temporal change in the prevalence of D614G in the Middle East and North Africa stratified by months of SARS-CoV-2 sequence collection.

### Maximum likelihood phyloegentic tree of MENA S gene sequences

To assess the possible presence of phylogenetic clusters in the MENA, ML analysis was conducted. The constructed ML tree showed a star-shaped pattern with short internal branches and long terminal branches (Figure 3). A total of 13 phylogenetic clusters (aLRT-SH ≥ 0.9) were determined; eight of which included sequences from a single MENA country and five clusters contained sequences collected in more than one MENA country (Appendix S2). Five clusters contained two sequences, and two large clusters were identified, each containing 26 MENA sequences. The highest percentage of clustering sequences was found in Iran (n = 3/5, 60.0%), followed by KSA (n = 39/149, 27.9%), and Tunisia (n = 2/8, 25.0%, Figure 4). The overall proportion of phylogenetic clustering was 15.4% (n = 85/553).

**Figure 3.**
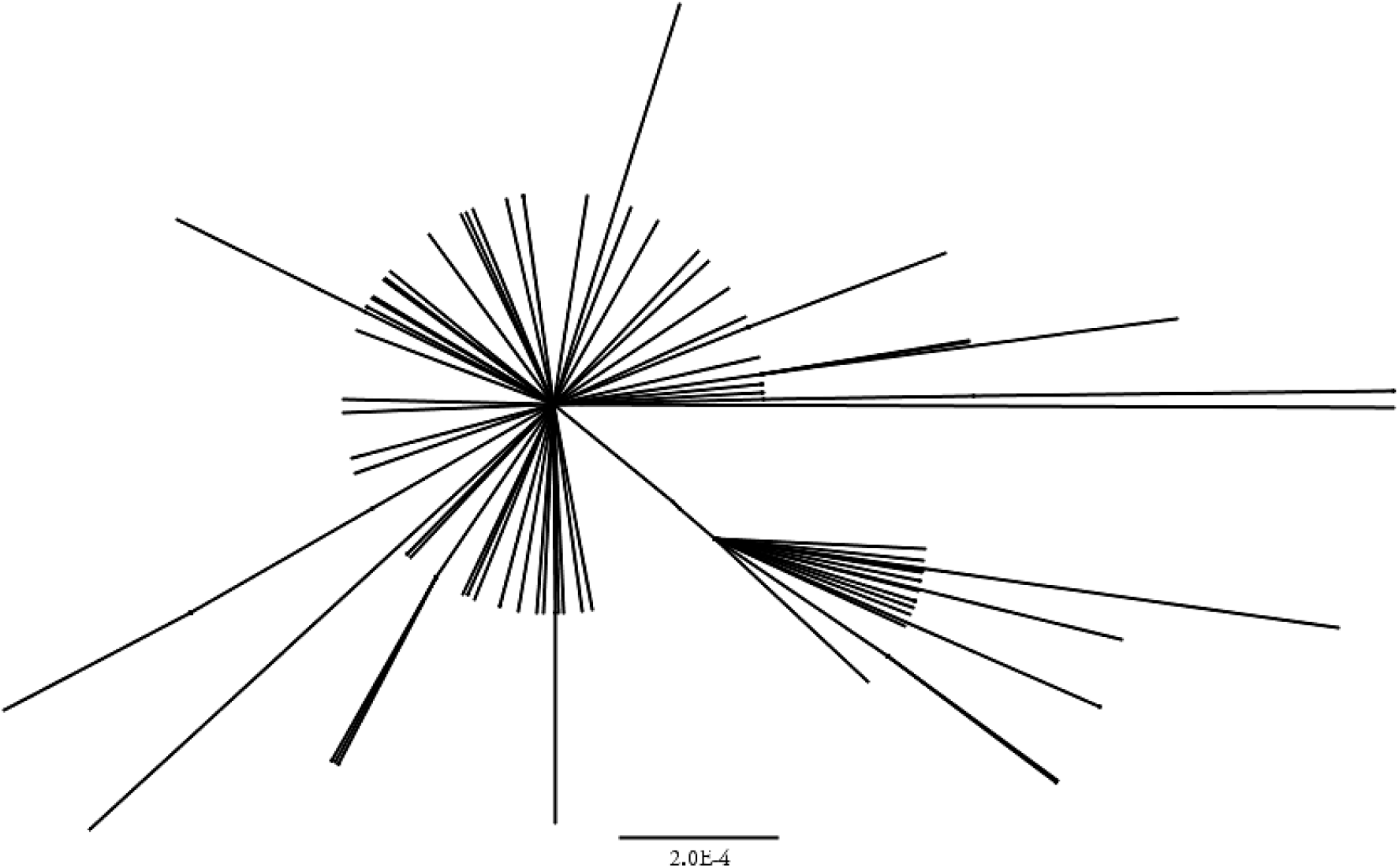
Maximum likelihood tree of the 533 Middle East and North Africa (MENA) spike (*S*) sequences showing a star-like shape.

**Figure 4.**
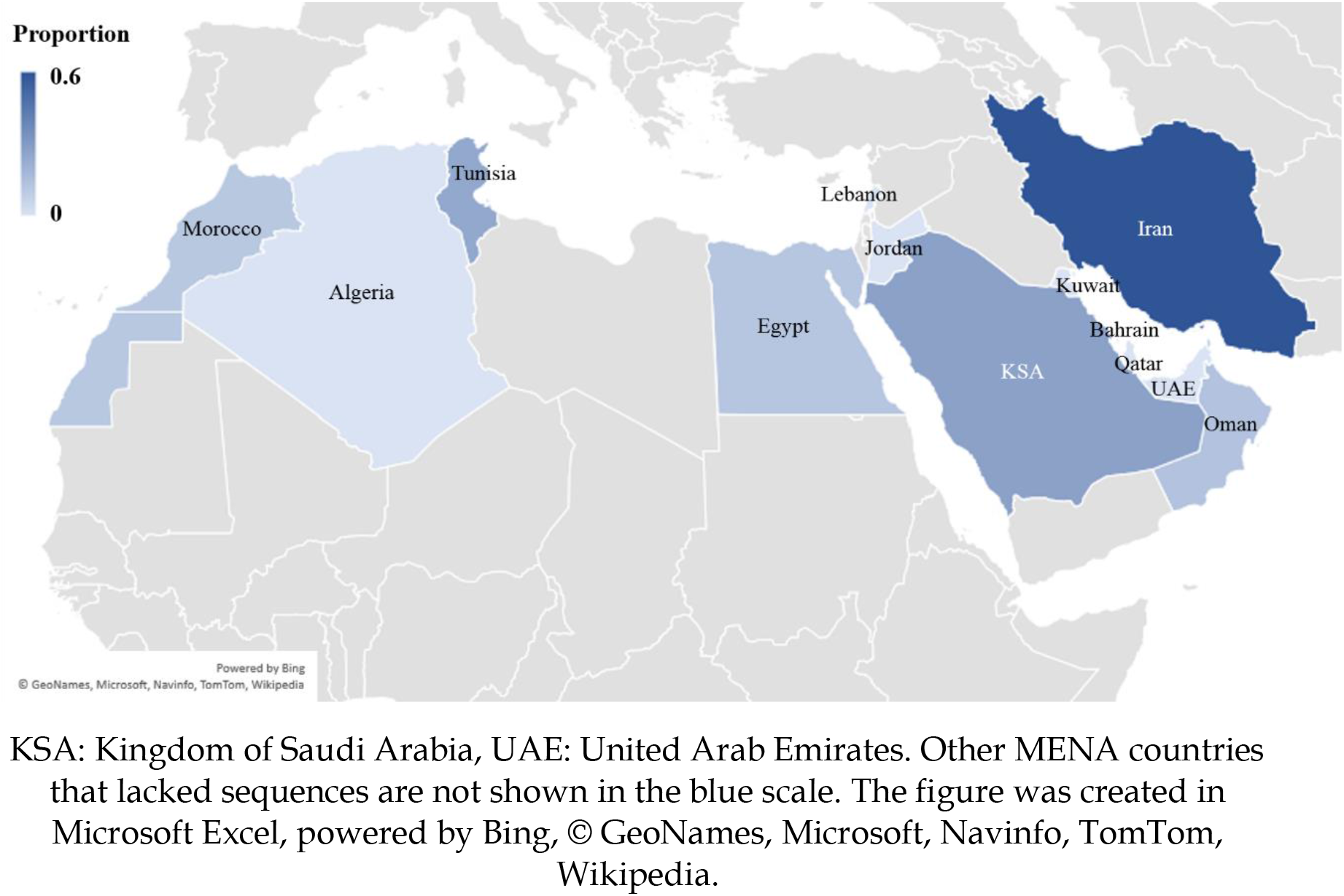
The Middle East and North Africa (MENA) map showing the proportion of phylogenetic clustering among the spike (*S*) sequences as inferred by maximum likelihood analysis.

### Bayesian analysis of the largest MENA phylogenetic clusters

Bayesian phylogenetic analysis was conducted on the two large clusters identified previously using the ML approach. One Egyptian sequence was removed from each cluster due to the lack of exact collection date. This resulted in analysis of two clusters, each containing 25 sequences. The first cluster contained 14 Saudi sequences, ten Omani sequences and a single Egyptian sequence, with a range of sequence collection between February 13 and May 11. The median estimate for tMRCA for this cluster having the D614G mutation was February 8, 2020 (95% highest posterior density interval [HPD]: October 19, 2019–February 13, 2020, Figure 5). For the second cluster (D614) with 20 Saudi sequences, three Egyptian sequences and two Tunisian sequences, the estimated median tMRCA was March 15, 2020 (95% HPD: February 21, 2020–March 15, 2020). The mean evolutionary rate estimated by molecular clock analysis was 6.46×10^−3^ substitutions/site/year (s/s/y) for the first cluster (95% HPD: 4.87×10^−3^ – 8.03×10^−3^s/s/y), and 6.50×10^−3^ s/s/y for the second cluster (95% HPD: 4.91×10^−3^ – 8.03×10^−3^s/s/y).

**Figure 5.**
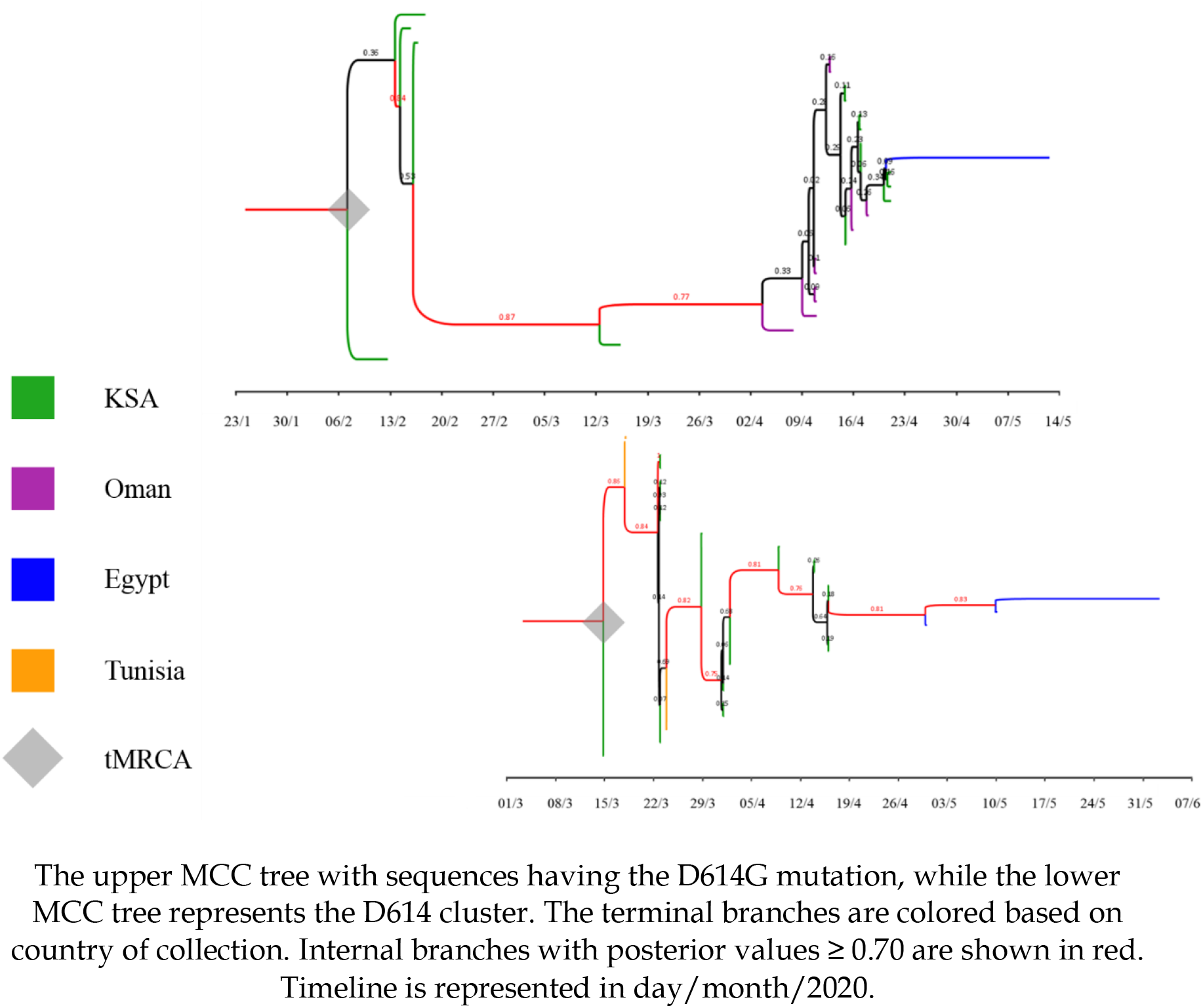
Maximum clade credibility (MCC) trees of the two large Middle East and North Africa (MENA) SARS-CoV-2 (Severe acute respiratory syndrome coronavirus 2) phylogenetic clusters.

## Discussion

In this study, phylogenetic analysis tools were utilized to assess origins, spread and mutations of SRAS-CoV-2 in the MENA. Phylogeny construction can help to formulate hypotheses regarding the spread of certain taxa having a common origin (Ciccozzi *et al*., 2019; Pybus and Rambaut, 2009). In addition, molecular clock analysis can help to establish a timeline for origins of monophyletic clades (Jenkins *et al*., 2002; Nasir and Caetano-Anolles, 2015). Phylogenetic analysis of the MENA *S* gene SARS-CoV-2 sequences showed a relatively low level of phylogenetic clustering (15%), which hints to a large number of virus introductions into the region. In addition, molecular clock analysis suggests an early introduction of the virus into the MENA which might have been circulating in the region from early February 2020 or even earlier, with subsequent spread into large networks of virus transmission. This estimate of an early virus introduction is supported by the close proximity in time of official reporting of confirmed COVID-19 cases in the region (Karamouzian and Madani, 2020).

In this study, no evidence of distinct SARS-CoV-2 genetic variants was found. Plausible explanations might be related to the use of sub-genomic part of the genome (the *S* gene) rather than utilizing the whole genome. The rationale behind selecting the *S* region was for two reasons: first, the variability of this region is expected to be higher than other parts of the genome (e.g. *RdRp*, where mutations are more costly) (Agostini *et al*., 2018; Shannon *et al*., 2020). Second, mutations in the *S* gene can have significant impact particularly for vaccine development and utility of neutralizing antibodies (Lokman *et al*., 2020). The absence of distinct SARS-CoV-2 genetic variants in this study does not provide a conclusive evidence of its genuine absence from the region. These two genetic variants (named L and S lineages) were reported previously, however, a recent report by MacLean *et al*. carefully discussed the potential pitfalls of such premature conclusions (MacLean *et al*., 2020; Tang *et al*., 2020).

For the estimated evolutionary rate of the two large MENA clusters identified in this study, we based the rate prior selection on the previous finding by (Giovanetti *et al*., 2020). This estimate appears higher than other estimates for SARS-CoV-2 and should be interpreted with caution based on our selection of a strong prior. However, the rate estimate might appear plausible, since it represents the *S* gene, rather than the whole genome. For ML analysis, the MENA sequences yielded a star-like phylogeny suggesting a recent growing epidemic (Colijn and Plazzotta, 2018).

The major result of this study was the demonstration of a temporal shift of SARS-CoV-2 from D614 into D614G variant, which dominated the most recent sequences collected in the region. Such trend was revealed at the global level by Korber *et al*., and our results indicated a similar pattern in the MENA (Korber *et al*., 2020). In the aforementioned comprehensive study, Korber *et al*. estimated the global prevalence of D614G at 71.0%, whereas our estimate in the MENA was 78.7%, which appears reasonable, bearing in mind the protracted duration of sequence collection in this study. The explanation for such an observation is most likely related to the association of D614G with a higher viral load and subsequent higher quantities of the virus shed by infected individuals, which increases the likelihood of infection by such a mutant, although an early founder effect of this variant cannot be ruled out (Deng *et al*., 2020; Farkas *et al*., 2020; Yurkovetskiy *et al*., 2020; Zhang *et al*., 2020). Whether this variant can have an effect on severity and outcome of COVID-19 is yet to be fully determined (Becerra-Flores and Cardozo, 2020; Eaaswarkhanth *et al*., 2020; Korber *et al*., 2020). This mutation appeared in all MENA countries, except in Qatar and Iran, which might be related to the low number of sequences from these two countries that were found in GISAID, and the early time of sequence collection (less than 10 sequences from each country were found, dating back to March, 2020). The emergence of D614G and its increasing prevalence have been reported by several published papers and preprints including a report from North Africa by Laamarti *et al*., albeit with a fewer number of sequences than the one analyzed in the current study (Gong *et al*., 2020; Kim *et al*., 2020b; Laamarti *et al*., 2020; Maitra *et al*., 2020).

Other mutations that were found in the study included Q677H (found only in Egypt), and L5F found in three different countries (Oman, Egypt, and Morocco). The L5F mutation is located in the signal peptide domain of the spike glycoprotein and might be related to recurring sequencing errors (Korber *et al*., 2020; N. De Maio, 2020). Nevertheless, its appearance in different studies warrants further investigation to determine its significance (Korber *et al*., 2020). The functional importance of Q677H has not been determined yet despite a previous report describing its occurrence (Kim *et al*., 2020a).

Limitations of this study should be clearly stated and taken into consideration. The most obvious caveat in the study was sampling bias. In spite of reporting COVID-19 in all MENA countries, the following countries did not have *S* sequences submitted to GISAID: Syria, Libya, Yemen, Sudan and Palestine (Iraq had partial sequences that did not include the *S* gene). In addition, bias was observed for timing of sequence collection. Furthermore, only two countries (Oman and KSA) had more than 100 sequences available for analysis. Another point that should be considered is related to the molecular clock analysis, where we used a strong informative prior which may have affected our tMRCA estimates for dating the origins of the two large phylogenetic clusters. Sequencing errors should also be taken into account, which can partly explain some sporadic mutations that were found in this study.

## Data Availability

The datasets analysed during the current study are available from the corresponding author on reasonable request and considering the terms of use by GISAID.

## Conclusions

In the current study, we demonstrated that the D614G variant of SARS-CoV-2 appears to be taking over COVID-19 epidemic in the MENA, similar to what have been reported in other regions around the globe. Local transmission of SARS-CoV-2 might have been established earlier than previously thought, and this illustrates the importance of vigilant surveillance in such conditions of outbreaks by novel viruses. The mutational patterns of SARS-CoV-2 should be closely monitored as the virus seems to be heading into an endemicity in the human population, particularly in relation to mutations’ potential impact on passive and active immunization.

## Supplementary Materials

Appendix S1: A complete list of the MENA SARS-CoV-2 sequence epi accession numbers that were analyzed in this study.

Appendix S2: Maximum likelihood tree of the 553 MENA *S* gene sequences.

## Author Contributions

Conceptualization, M.S. and A.M.; methodology, M.S., N.A.A., D.D., F.G.B and A.M.; software, M.S.; validation, M.S.; formal analysis, M.S.; investigation, M.S., N.A.A., D.D., F.G.B and A.M.; data curation, M.S.; writing— original draft preparation, M.S.; writing—review and editing, M.S., N.A.A., D.D., F.G.B and A.M.; visualization, M.S.; supervision, M.S and A.M.; project administration, M.S. All authors have read and agreed to the published version of the manuscript.

## Acknowledgments

None.

